# Machine learning-based CT radiomics model for predicting hospital stay in patients with pneumonia associated with SARS-CoV-2 infection: A multicenter study

**DOI:** 10.1101/2020.02.29.20029603

**Authors:** Xiaolong Qi, Zicheng Jiang, Qian Yu, Chuxiao Shao, Hongguang Zhang, Hongmei Yue, Baoyi Ma, Yuancheng Wang, Chuan Liu, Xiangpan Meng, Shan Huang, Jitao Wang, Dan Xu, Junqiang Lei, Guanghang Xie, Huihong Huang, Jie Yang, Jiansong Ji, Hongqiu Pan, Shengqiang Zou, Shenghong Ju

## Abstract

**Objectives:** To develop and test machine learning-based CT radiomics models for predicting hospital stay in patients with pneumonia associated with SARS-CoV-2 infection.

**Design:** Cross-sectional

**Setting:** Multicenter

**Participants:** A total of 52 patients with laboratory-confirmed SARS-CoV-2 infection and their initial CT images were enrolled from 5 designated hospitals in Ankang, Lishui, Zhenjiang, Lanzhou, and Linxia between January 23, 2020 and February 8, 2020. As of February 20, patients remained in hospital or with non-findings in CT were excluded. Therefore, 31 patients with 72 lesion segments were included in the final analysis.

**Intervention:** CT radiomics models based on logistic regression (LR) and random forest (RF) were developed on features extracted from pneumonia lesions in training and inter-validation datasets. The predictive performance was further evaluated in test dataset on lung lobe- and patients-level.

**Main outcomes:** Short-term hospital stay (≤10 days) and long-term hospital stay (>10 days).

**Results:** The CT radiomics models based on 6 second-order features were effective in discriminating short- and long-term hospital stay in patients with pneumonia associated with SARS-CoV-2 infection, with areas under the curves of 0.97 (95%CI 0.83-1.0) and 0.92 (95%CI 0.67-1.0) by LR and RF, respectively, in the test dataset. The LR model showed a sensitivity and specificity of 1.0 and 0.89, and the RF model showed similar performance with sensitivity and specificity of 0.75 and 1.0 in test dataset.

**Conclusions:** The machine learning-based CT radiomics models showed feasibility and accuracy for predicting hospital stay in patients with pneumonia associated with SARS-CoV-2 infection.

## Introduction

The coronavirus disease 2019 (COVID-19) from Wuhan, China has become a global challenge since the December 2019.^1-3^ Clinical characteristics of patients with severe acute respiratory syndrome coronavirus 2 (SARS-CoV-2) infection have been reported, and the median hospital stay of forty-seven discharged patients was 10 days.^1^ Studies showed that patients’ condition in Wuhan worsens on the 10th day after illness onset.^2^ However, patients with symptoms longer than 10 days outside Wuhan were less severe than those in Wuhan.^3^ Therefore, the hospital stay in patients with SARS-CoV-2 infection is one of the prognostic indicators, and its non-invasive predicting tool is important for assessing the patients’ clinical outcome.

Chest CT is recommended as a routine test in the diagnoses and monitoring of COVID-19 since ground-glass opacities and consolidation are the most relative imaging features in pneumonia associated with SARS-CoV-2 infection.^4-6^ On the basis of our previous work utilizing quantitative CT for COVID-19,^7^ we hypothesized that high-throughout information hidden behind CT images^8^ had potential in discriminating the hospital stay. The study aimed to develop and test machine learning-based CT radiomics models for predicting hospital stay in patients with pneumonia associated with SARS-CoV-2 infection.

## Methods

### Study population

The multicenter study was conducted according to principles of the Declaration of Helsinki and approved by all institutional review board. The need for written informed consent from the participants was waived. Patients with laboratory-confirmed SARS-CoV-2 infection and their initial CT images were enrolled from 5 designated hospitals between January 23, 2020 and February 8, 2020, with final follow-up on February 20, 2020 (**Figure 1**). Most patients received antiviral treatment with interferon inhalation, lopinavir and ritonavir, combined with probiotics. Patients were discharged once the results of two real-time fluorescence polymerase-chain-reaction tests taken 24 hours apart were negative for SARS-CoV-2 antigens. Patients without pneumonia findings or those remained in hospital were excluded. Sample size consideration was shown in supplementary file. In this study, the optimal cut-off value of hospital stay was determined to be 10 days based on previous studies^1-3^, by which patients were classified into short-term hospital stay (≤10 days) and long-term hospital stay (>10 days).

**Figure 1.**
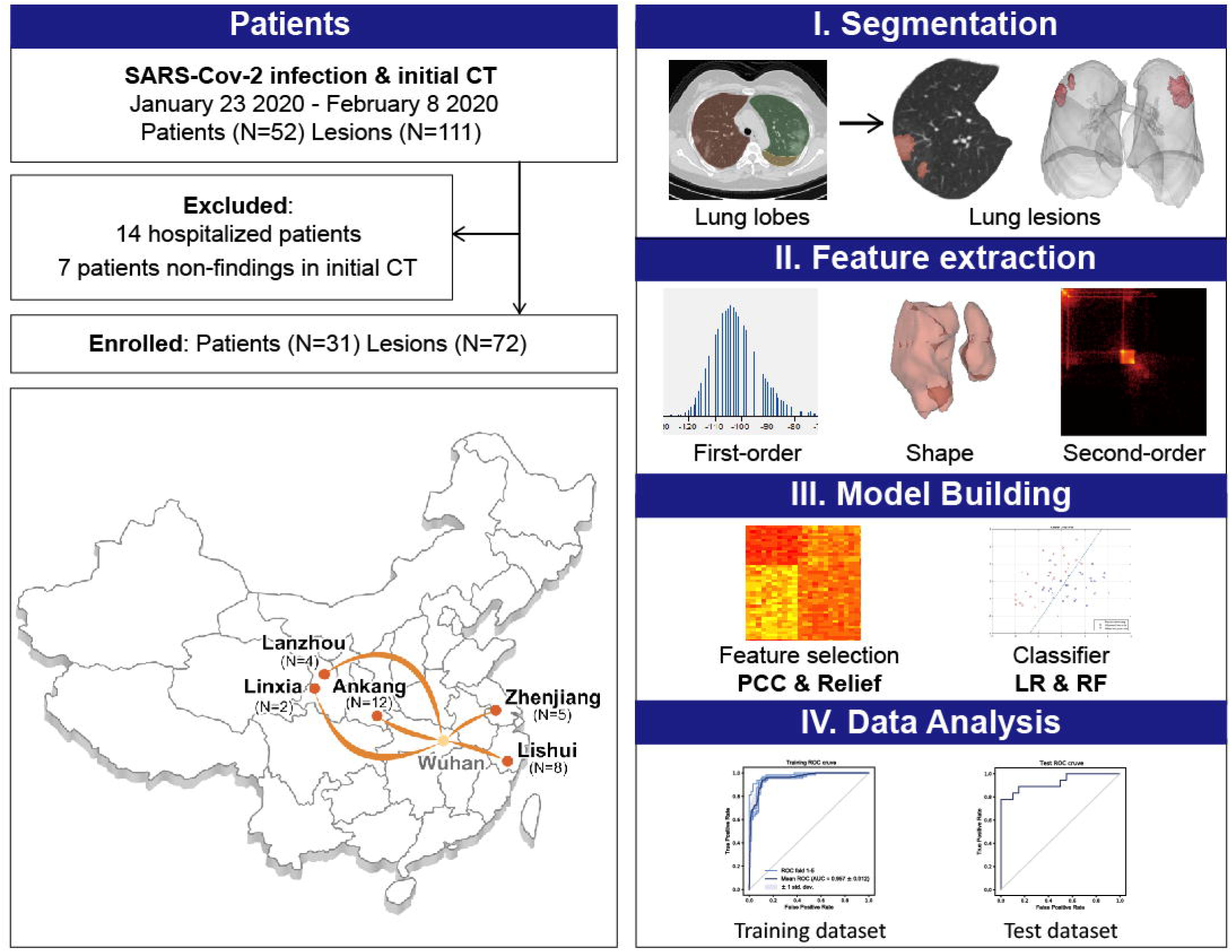
Flow chart of study population and model building. PCC, Pearson correlation coefficient; LR, logistic regression; RF, random forest.

### CT radiomics features and model building

The pipeline of radiomics model was shown (**Figure 1**), and features extraction and model building were performed on lung lobe-level. Images preprocessing was shown in supplementary file. Images containing lesions were segmented using Python (3.6, https://www.python.org) and 3Dslicer (version 4.10.0; https://www.slicer.org/) with two steps. First, lung lobes of each patient were segmented automatically using algorithms based on U-net^9^, and results were checked and modified by one radiologist (Q.Y). Next, lesions in each lung lobe were labeled semi-automatically using serval seeds placed within lesion region to generate the contours. Three radiologists (S.H, Q.Y and X.P.M) evaluated segments of each lesion and reached a consensus. All imaging processes were blinded to clinical data.

In total, 1218 features were calculated per lesion patch. First-order, shape and second-order features were extracted from original images and wavelet filter applied images using pyradiomics.^10^ Two supervised learning algorithms, logistic regression (LR) and random forest (RF), were used to build the model and verify the robustness of features (supplementary file).^11^ We applied 5-fold cross-validation on the training dataset to prove model performance.

### Model performance

The cutoff point was defined on receiver operating characteristic (ROC) curves of training data by maximizing the sum of sensitivity and specificity. The model performance was evaluated using test dataset on lung lobe-level. Areas under the ROC curve (AUC), sensitivities, specificities, positive predictive value (PPV), and negative predictive value (NPV) were recorded. On patient-level, one was defined as long-term hospital stay once more than one lesion of lung lobe was labeled as long-term stay lesion, if not, as short-term hospital stay.

### Statistical analysis

Categorical data were expressed in numbers (percentages), and continuous variables as median (interquartile range). Demographic and clinical characteristics of patients were assessed using Chi square test (Fisher’s exact test as appropriate) for categorical variables, and Mann-Whitney test for continuous variables in SPSS (version 22.0. IBM Crop. Armonk, NY, USA). Feature selection and model building were implemented with FeAture Explorer (FAE, v0.2.5, https://github.com/salan668/FAE) on Python (3.6.8, https://www.python.org/)). Test values like areas under the receiver operating characteristic curves (95% confidence interval), sensitivity, specificity was calculated in SPSS and Python. A P-value < 0.05 was considered statistically significant.

### Patient and Public Involvement

This cross-sectional study did not involve patients in study design, outcome measures, or writing or editing of this study.

## Results

### Patient characteristics

A total of 52 patients with laboratory-confirmed SARS-CoV-2 infection and initial CT images were enrolled from 5 designated hospitals in Ankang, Lishui, Zhenjiang, Lanzhou, and Linxia, China. As of February 20, 14 patients were still hospitalized, and 7 patients had non-findings in CT images. Therefore, 31 patients with 72 lesion segments were included in the final analysis. The training and inter-validation cohort comprised 26 patients (12 from Ankang, 8 from Lishui, 4 from Lanzhou, and 2 from Linxia) with 59 lesion segments, and test cohort comprised 5 patients from Zhenjiang with 13 lesion segments. The median age was 38.00 (interquartile range, 26.00-47.00) years and 17 (57%) were male. Comorbidities, symptoms and laboratory findings at admission were summarized in **Table 1**.

**Table 1.** Baseline characteristics.

| Characteristics | All | Short-term stay (≤10days) | Long-term stay (>10days) | P value |
| --- | --- | --- | --- | --- |
| Age, median (interquartile range [IQR]), years | 38.00 | 32.00 (26.00-46.50) | 41.00 (26.00-47.00) | 0.72 |
| Gender, No. (%), male | 17 (55%) | 2 (25%) | 15 (65%) | 0.10 |
| Comorbidities, No. (%) | 3 (10%) | 1 (13%) | 2 (9%) | 1 |
| Hypertension, No. (%) | 2 (6%) | 1 (13%) | 1 (4%) |  |
| History of cancer, No. (%) | 2 (6%) | 0 (0%) | 2 (9%) |  |
| Symptoms |  |  |  |  |
| Fever, No. (%) | 22 (71%) | 5 (63%) | 17 (74%) |  |
| Myalgia, No. (%) | 6 (19%) | 2 (25%) | 4 (17%) |  |
| Diarrhea, No. (%) | 2 (6%) | 1 (13%) | 1 (4%) |  |
| Headache, No. (%) | 11 (35%) | 3 (38%) | 8 (35%) |  |
| Cough, No. (%) | 25 (81%) | 7 (88%) | 18 (78%) |  |
| Laboratory findings |  |  |  |  |
| Blood leukocyte count, median (IQR), ×10<sup>9</sup>/L | 4.42 | 5.09 (3.70-6.89) | 4.30 (3.56-4.66) | 0.14 |
| Neutrophil count, median (IQR), ×10<sup>9</sup>/L | 2.83 | 3.25 (2.45-4.19) | 2.80 (1.90-3.30) | 0.10 |
| Neutrophil percentage, median (IQR), % | 62.30 | 63.15 (61.63-71.80) | 61.30 (52.90-65.50) | 0.32 |
| Lymphocyte count, median (IQR), $\times 10^9/L$ | 1.30 | 1.38 (1.16-1.93) | 1.22 (0.91-1.40) | 0.17 |
| Lymphocyte percentage, median (IQR), % | 29.00 | 26.70 (21.63-29.45) | 31.90 (23.2-36.10) | 0.30 |

### Performance of CT radiomics model

The CT radiomics model, based on 6 features (**supplementary Table1**), showed the highest AUC on the training and inter-validation dataset. The performance of modeling using LR and RF methods was shown in **Figure 2**. On lung lobe-level, models using LR method significantly distinguished short- and long-term hospital stay (In training and inter-validation datasets, cut-off value 0.31, AUC 0.94 (95%CI 0.92-0.97), sensitivity 1.0, specificity 0.87, NPV 1.0, and PPV 0.88; In test dataset, AUC 0.97 (95%CI 0.83-1.0), sensitivity 1.0, specificity 0.89, NPV 1.0, and PPV 0.8). Besides, models using RF method obtained satisfied results (In training and inter-validation datasets, cut-off value 0.68, AUC 1.0 (95%CI 1.0-1.0), sensitivity 1.0, specificity 1.0, NPV 1.0, and PPV 1.0; In test dataset, AUC 0.92 (95%CI 0.67-1.0), sensitivity 0.75, specificity 1.0, NPV 0.9, and PPV 1.0).

**Figure 2.**
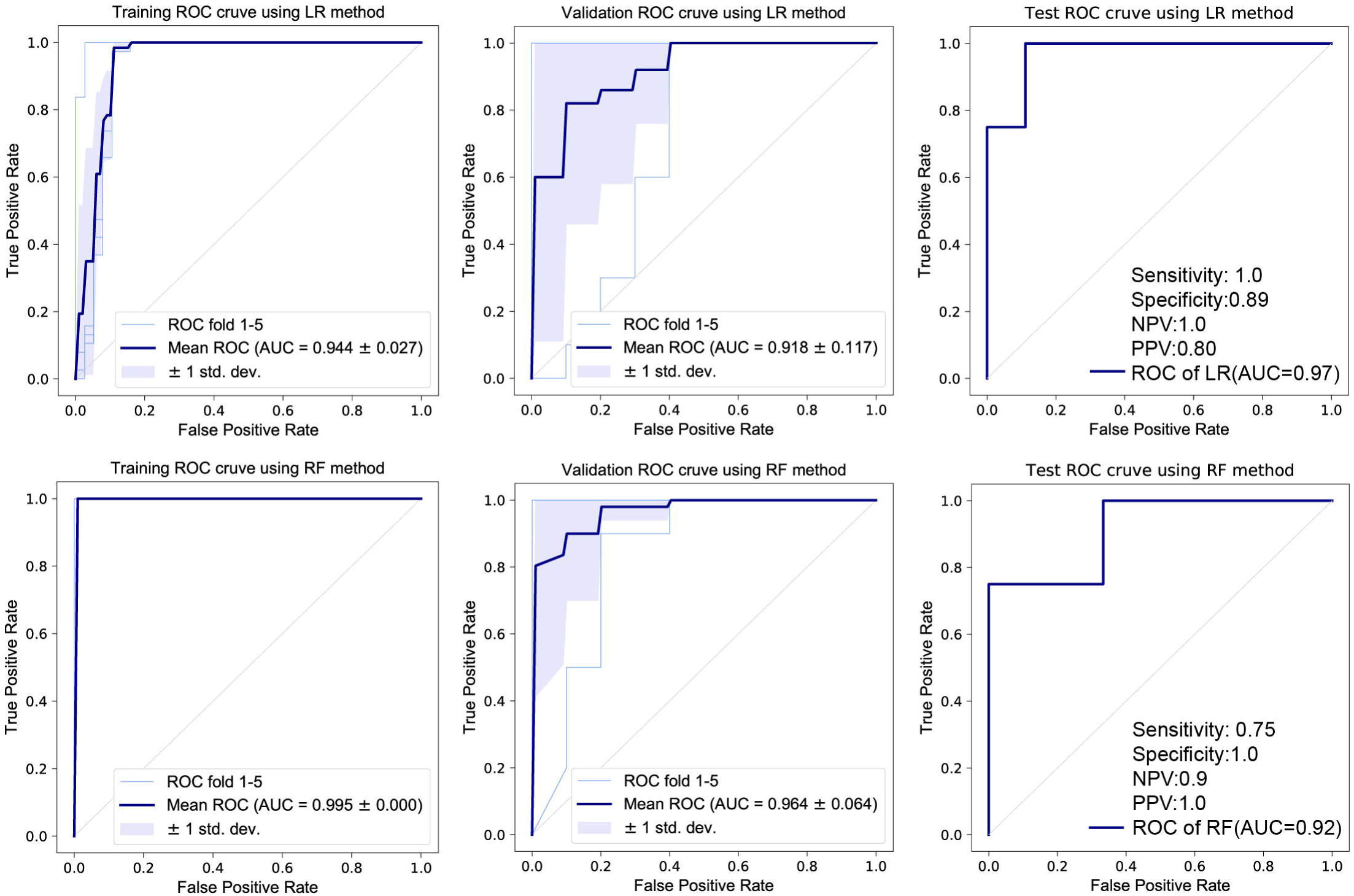
The predictive performance of CT radiomics models in training, inter-validation and test datasets. ROC, receiver operating characteristic; AUC, areas under the ROC curves; LR, logistic regression; RF, random forest; PPV, positive predictive value; NPV, negative predictive value.

On patients-level, in training and inter-validation datasets, 6 of 6 patients were correctly classified as short-term stay by both models, and 20 of 20, 16 of 20 patients were correctly identified as long-term stay by RF and LR models, respectively. In test dataset, 1 of 2 patients was correctly classified as short-term stay, and 3 of 3 were correctly identified as long-term stay by RF and LR models.

As of February 28, we followed up for a prospective dataset of 6 newly discharged patients with 24 lesions from designated hospitals (Comorbidities, symptoms and laboratory findings were described in supplementary Table 2). All patients were correctly recognized as long-term stay using both RF and LR models developed on raw 52 patients.

## Discussion

In this study, machine learning-based CT radiomics models were developed and tested for predicting hospital stay in patients with pneumonia associated with SARS-CoV-2 infection. CT radiomics features hidden within lesions, ground-glass opacities and consolidation, were extracted, and machine-learning models demonstrated robust performance using multicenter cohorts for training and inter-validation, and an independent dataset and a prospective dataset for test.

Though there were slightly differences in CT scan parameters among centers, key features included in models were second-order, and focused on distribution, correlation and variance in gray level intensities, which described the relationship between voxels and hold quantitative information on the spatial heterogeneity of pneumonia lesions.^11,12^ Compared with first-order features, second-order features were not sensitive to absolute value and thus more robust. Moreover, the models showed satisfied AUCs more than 90% on both training and test process, which indicated that the models could be applied in a general situation.

Similarity in AUCs, sensitivity and specificity for RF and LR models also demonstrated the robustness, according to prior study that classification method showed most dominant in variability of model.^11^

The study was limited by small sample size. The percentage of short-term hospital stay is low in our multicenter cohorts, and semi-automated lesion segmentation might result in selection bias. A larger prospective multicenter cohort is needed to tune and test the machine learning-based CT radiomics models.

In summary, the machine learning-based CT radiomics models showed feasibility and accuracy for predicting hospital stay in patients with pneumonia associated with SARS-CoV-2 infection.

## Data Availability

The imaging or algorithm data used in this study are available upon request.

## Summary boxes

### What is already known on this topic

The coronavirus disease 2019 (COVID-19) from Wuhan, China has become a global challenge since the December 2019. The median hospital stay of discharged patients with severe acute respiratory syndrome coronavirus 2 (SARS-CoV-2) infection was reported as 10 days.

The condition of patients in Wuhan worsens on the 10th day after illness onset. However, patients with symptoms longer than 10 days outside Wuhan were less severe than those in Wuhan.

The hospital stay is one of the prognostic indicators, and its non-invasive predicting model based on CT radiomics features is important for assessing the patients’ clinical outcome.

### What this study adds

In the multicenter study with patients from 5 designated hospitals in China, the CT radiomics models using logistic regression and random forest method showed satisfied diagnostic performance with sensitivity of 1.0 and 0.75, specificity of 0.89 and 1.0, respectively, in independent test dataset.

The machine learning-based CT radiomics models showed feasibility and accuracy for predicting hospital stay in patients with pneumonia associated with SARS-CoV-2 infection.

